# Biallelic *CYB5A* disruptions in 46,XY Disorder of Sex Development: Identification and Characterization of a Novel Deep Intronic Variant

**DOI:** 10.64898/2026.05.05.26352416

**Authors:** Shirin Moradifard, Thanh Nha Uyen Le, Nguyen Thu Ha, Vu Chi Dung, Bui Phuong Thao, Vincent Harley

**Affiliations:** Hudson Institute of Medical Research, Melbourne, Australia; Dept. of Molecular and Translational Science, Monash University, Melbourne, Australia; National Children Hospital, Vietnam

**Keywords:** CYB5A, 46, XY DSD, WGS

## Abstract

**Background:** The diagnostic yield for 46,XY disorders of sex development (DSD) remains limited. Whole-genome sequencing (WGS) improves detection of both coding and non-coding variants that may be missed by routine testing. Cytochrome b5, encoded by CYB5A, is an essential co-factor for CYP17A1-mediated 17,20-lyase activity. We report on WGS on a Vietnamese family with 46,XY DSD with two siblings presenting with female external genitalia.

**Methods:** Clinical assessment and hormone profiling were conducted. WGS was conducted on peripheral blood DNA, in two affected siblings followed by variant annotation and ACMG-based classification. A minigene RNA splicing assay in HEK293 cells was used to evaluate the functional impact of the *CYB5A* intronic variant.

**Results:** The patient’s hormone profile showed low testosterone and estradiol. WGS identified compound-heterozygous *CYB5A* variants: a paternally inherited missense variant (p.Val34Glu, likely pathogenic) and a maternally inherited deep intronic deletion (c.129+862_129+863del) for which SpliceAI predicted aberrant splicing. Minigene assays confirmed that the intronic deletion creates cryptic splice sites, resulting in pseudoexon inclusion and a premature stop codon, consistent with nonsense-mediated decay. The intronic variant meets ACMG criteria for pathogenicity.

**Conclusion:** This family expands the spectrum of *CYB5A*-related DSD and demonstrates that compound-heterozygous variants, including deep intronic defects, can lead to a disruption in 17,20-lyase activity. These findings highlight the importance of WGS and functional assays for identifying clinically relevant non-coding variants in DSD.

## Introduction

Differences of sex development (DSD) is a group of congenital disorders relating to the aberrant development of chromosomal, gonadal or external genitalia (1). Although whole exome sequencing (WES) is used in clinical practice, a molecular diagnosis is only obtained in 30% to 50% of cases. This results in a substantial diagnostic gap that affects patient counseling and long-term care (2, 3). This diagnostic challenge largely reflects the biochemical overlap among steroidogenic disorders, where defects in different enzymes can give rise to similar hormonal profiles.

Cytochrome P450 family 17 subfamily A member 1 (*CYP17A1*) gene plays a crucial role in human steroidogenesis, by encoding 17α-hydroxylase and 17,20-lyase enzymes. 17α-hydroxylase converts pregnenolone and progesterone into 17α-hydroxypregnenolone and 17α-hydroxyprogesterone (17OHP), respectively. However, 17,20-lyase activity catalyzes the conversion of 17α-hydroxypregnenolone and 17OHP into dehydroepiandrosterone (DHEA) and androstenedione, respectively. These intermediates are crucial precursors for producing glucocorticoids like cortisol and sex steroids (4, 5).

Deficiency in 17 α-hydroxylase (17OHD) is a rare inherited disorder with the frequency of 1% in congenital adrenal hyperplasia (CAH) cases and it results from a mutation in the *CYP17A1* gene (4, 5). Lack of the 17α-hydroxylase enzyme results in decreased cortisol and androgen levels, which causes an increase in adrenocorticotropic hormone (ACTH), gonadotropins, and 11-deoxycorticosterone production (5). Androgen deficiency in males’ results in ambiguous or female external genitalia. Female patients have normal genitalia, but because of estrogen deficiency during adolescence, they experience delayed sexual development and primary amenorrhea (6). Total 17OHP deficiency is well known and usually manifests with cortisol deficiency, mineralocorticoid excess, hypertension, hypokalemia, and disrupted sexual development. (7, 8).

In contrast, isolated 17,20-lyase deficiency is rare and less obvious. Here, cortisol levels remain normal, but androgen production is specifically compromised. This specific impairment usually occurs due to malfunction of accessory proteins such as P450 oxidoreductase and cytochrome b5 (*CYB5A*), rather than mutations directly in *CYP17A1* (6, 9-12).

CYB5A is a small hemoprotein that exists in two isoforms produced by alternative splicing: a soluble 98 amino acid form found in reticulocytes and adrenal tissue, and a membrane-bound 134 amino acid form present in various human tissues. A deficiency in *CYB5A* is a known cause of autosomal recessive congenital methemoglobinemia (13-15). While the role of CYB5A is well known, only three variants have been classified as pathogenic or likely pathogenic. This small number may partly result from the widespread use of WES and targeted gene panels, which mainly focus on coding regions and canonical splice sites. Consequently, deep intronic and other non-coding variants that affect splicing might be overlooked.

In this study, we reported two siblings with 46,XY DSD who have compound heterozygous variants in *CYB5A*. These include a novel missense variant and a novel deep intronic deletion. A minigene splicing assay demonstrated that the intronic variant activates a cryptic splice site, disrupting normal *CYB5A* transcript processing. These results broaden the known mutation types in *CYB5A*, underscore the diagnostic importance of WGS in identifying non-coding pathogenic variants, and highlight the value of combining steroid metabolomics with functional genomics in DSD assessment.

## Materials and Methods

### Patients

The patients included in this study were two siblings presenting with 46,XY DSD, recruited from the National Children’s Hospital in Vietnam. Both individuals were born to unaffected parents. Written informed consent for genetic testing, use of clinical data, and publication was obtained from their parents. The study was approved by the Monash Health Human Research Ethics Committee and conducted in accordance with institutional and national ethical guidelines.

### Whole Genome Sequencing (WGS) and Variant Analysis

#### DNA Extraction and Quality Control

Peripheral blood was collected from both affected siblings and their parents for genomic analysis. Genomic DNA (gDNA) was extracted using the Promega kit according to the manufacturer’s instructions. Initial quality assessment was performed via NanoDrop. Before library preparation, sample integrity was further verified at the sequencing facility (BGI) using a Qubit Fluorometer. All samples passed stringent quality control criteria, maintaining an OD 260/280 > 1.8 and a concentration exceeding 28 ng/µL.

### Library Construction, Sequencing, and Data Quality

Genomic DNA from each individual was submitted to BGI for WGS. Short-insert libraries (800 bp) were prepared according to the manufacturer’s protocols and sequenced on the DNBSEQ™ platform using a paired-end 150 bp (PE150) strategy. Raw reads were processed to remove adapters and low-quality bases, generating clean data encoded in Phred+33 format. High-quality sequencing metrics were obtained for both siblings.

### Bioinformatics Analysis, Variant calling and Prioritization

Raw WGS reads were assessed with FastQC, and high-quality reads were aligned to the hg19 reference genome using BWA-MEM (16, 17). Post-alignment quality metrics were evaluated with Qualimap. GATK was used as the variant-calling pipeline (18), followed by normalization with bcftools and annotation with VEP (19). For downstream analysis, a minor allele frequency (MAF) threshold of <0.001 was applied using population databases such as gnomAD v4.1.0, and variants were further integrated into a unified database using GEMINI, which incorporates diverse annotation sources (e.g., dbSNP, ENCODE, UCSC, ClinVar, KEGG) to support interpretation and classification based on inheritance patterns. Variants were additionally evaluated using in-silico prediction tools including CADD, REVEL, and SpliceAI (Δscore > 0.2) (20-22), and prioritized according to allele frequency, predicted pathogenicity, inheritance pattern, and relevance to DSD genes. Variant classification was performed according to the (23).

### Sanger Sequencing-Variant Confirmation

The identified variants were confirmed in the siblings and their parents using Sanger sequencing. PCR amplification was performed with GoTaq® Green Master Mix (Promega), and the resulting amplicons were sequenced at Monash Genomics & Bioinformatics Platform with ABI 3930XL automated genetic analyzer (Applied Biosystems, Thermo Fisher Scientific). Primers were designed using Primer3.0, with the following sequences: forward 5′-GAACCGAGATGGCAG AGCAG-3′, and reverse 5′-CACCAAAAGCGCCCCTTAGC-3′.

### Minigene Splicing Assays

A minigene splicing assay was performed to investigate the functional effect of the deep intronic *CYB5A* variant NM_148923 c.129+862_129+863del. Genomic DNA from the mother, who is a heterozygous carrier of the variant, was used as the template to ensure amplification of both wild-type and mutant alleles. The pSPL3-*ABCC8*-9-10 vector served as the cloning backbone (24). Exon 9 and its flanking intronic regions were removed from the vector and replaced with *CYB5A* exon 1 together with 1082 bp of intron 1 immediately upstream of the endogenous splice junction for exon 10, thereby reconstructing the native *CYB5A* intron–exon context surrounding the predicted splice-altering region. The *CYB5A* fragment was amplified using primers containing EcoRI and PstI restriction sites (forward 5′-GATCACCAGAATTCATGGCAGAGCAGTCGGACGAG-3′ and reverse 5′-TACTCCTGCAGTGTAAAAAATTCCCATCACTAAACC-3′) and cloned into the pSPL3-*ABCC8*-9-10 vector using standard restriction–ligation methods. Because the maternal DNA contained both alleles, individual bacterial colonies were screened to isolate constructs carrying either the wild-type or mutant insert. Nanopore sequencing was used to confirm the integrity and identity of both *CYB5A* wild-type and mutant inserts within the vector.

Following construct validation, pSPL3-*ABCC8*-9-10 (splicing control), *CYB5A*-WT, and *CYB5A*-c.129+862_129+863del minigenes were transfected into HEK293T cells. A total of 80,000 cells were seeded per well in 6-well plates, and transfections were performed using Lipofectamine™ 2000 (Thermo Fisher Scientific) according to the manufacturer’s protocol. Cells were harvested 48 hours post-transfection. Total RNA was extracted using the RNeasy RNA Purification Kit (Qiagen), and cDNA synthesis was carried out with the QuantiTect Reverse Transcription Kit (Qiagen). Splicing products were amplified using vector-specific primers targeting exon 9–exon 10 of the pSPL3-*ABCC8*-9-10 backbone and *CYB5A*-specific primers spanning exon 1–exon 10 to distinguish wild-type and mutant transcript isoforms.

Following cDNA synthesis, RT-PCR was performed to assess transcript structure using primer sets specific to the minigene backbone and the *CYB5A* insert. For the pSPL3-*ABCC8*-9-10 control construct, amplification was carried out using an exon 9 forward primer (5′-TGGGTGTGATTCTCCTCTAC-3′) and an exon 10 reverse primer (5′-GGAGATGGAGGTATAGATGG-3′). For *CYB5A* constructs, transcript amplification employed a *CYB5A* exon 1 forward primer (5′-CTAGAGGAGATTCAGAAGCAC-3′) together with the same exon 10 reverse primer to distinguish wild-type and mutant splicing products. RT-PCR products were resolved on a 1.5% agarose gel, and bands of the expected sizes were excised and purified. Purified amplicons were submitted for Sanger sequencing to validate splice junctions and confirm transcript identity.

## Results

### Clinical Presentation

Clinical and biochemical data were collected from two siblings with 46,XY DSD at their initial presentation. Sibling 1 (proband 1) is a phenotypic female in the 6–10 year age range with a confirmed 46,XY karyotype. She presented with female-typical external genitalia and bilateral inguinal cryptorchidism. Pelvic ultrasound demonstrated the absence of Müllerian structures. At the time of assessment, her height was 119.5 cm and weight was 23.6 kg. Sibling 2 (proband 2) is a phenotypic female in the 1–5 year age range who also presented with 46,XY DSD and female-typical external genitalia. Clinical examination identified a urogenital sinus.

### Endocrine findings

Baseline endocrine evaluation in Sibling 1 (proband 1) showed markedly low serum testosterone (<0.24 nmol/L; reference <0.867 nmol/L) with 17-hydroxyprogesterone (17-OHP) 0.798 nmol/L (reference 0.3–5.2 nmol/L). Estradiol was 18.4 pmol/L (reference <43.3–90.7 pmol/L), and gonadotropins were within the normal prepubertal range (LH 0.3 IU/L; FSH 4.16 IU/L). Sibling 2 (proband 2) demonstrated a similar hormonal profile, with LH 0.93 IU/L (reference <0.1– 3.41 IU/L), FSH 1.61 IU/L (reference 1.12–9.51 IU/L), estradiol 18.4 pmol/L (reference <43.3– 90.7 pmol/L), and testosterone <0.24 nmol/L (reference <0.867 nmol/L). 17-hydroxyprogesterone was 2.97 nmol/L (reference 0.3–5.2 nmol/L).

### Genetic findings

Phenotype–genotype correlation and comprehensive database filtering identified compound heterozygous variants in *CYB5A* in both affected siblings. The paternally inherited missense variant NM_148923 c.101T>A (p.Val34Glu) showed a REVEL score of 0.931, supporting a deleterious effect. The maternally inherited deep intronic deletion c.129+862_129+863del was predicted by SpliceAI to create a cryptic donor gain site (score 0.68) and a low-level acceptor gain site (score 0.18), consistent with potential splice disruption. Both siblings carried both variants. All remaining variants were either located in genes unrelated to the clinical presentation or were present in heterozygous carrier states without phenotype correlation. Neither *CYB5A* variant has been reported in gnomAD v4.1.0 or ClinVar. Figure 1 shows the Sanger sequencing results confirming the inheritance of both variants from the parents.

**Figure 1:**
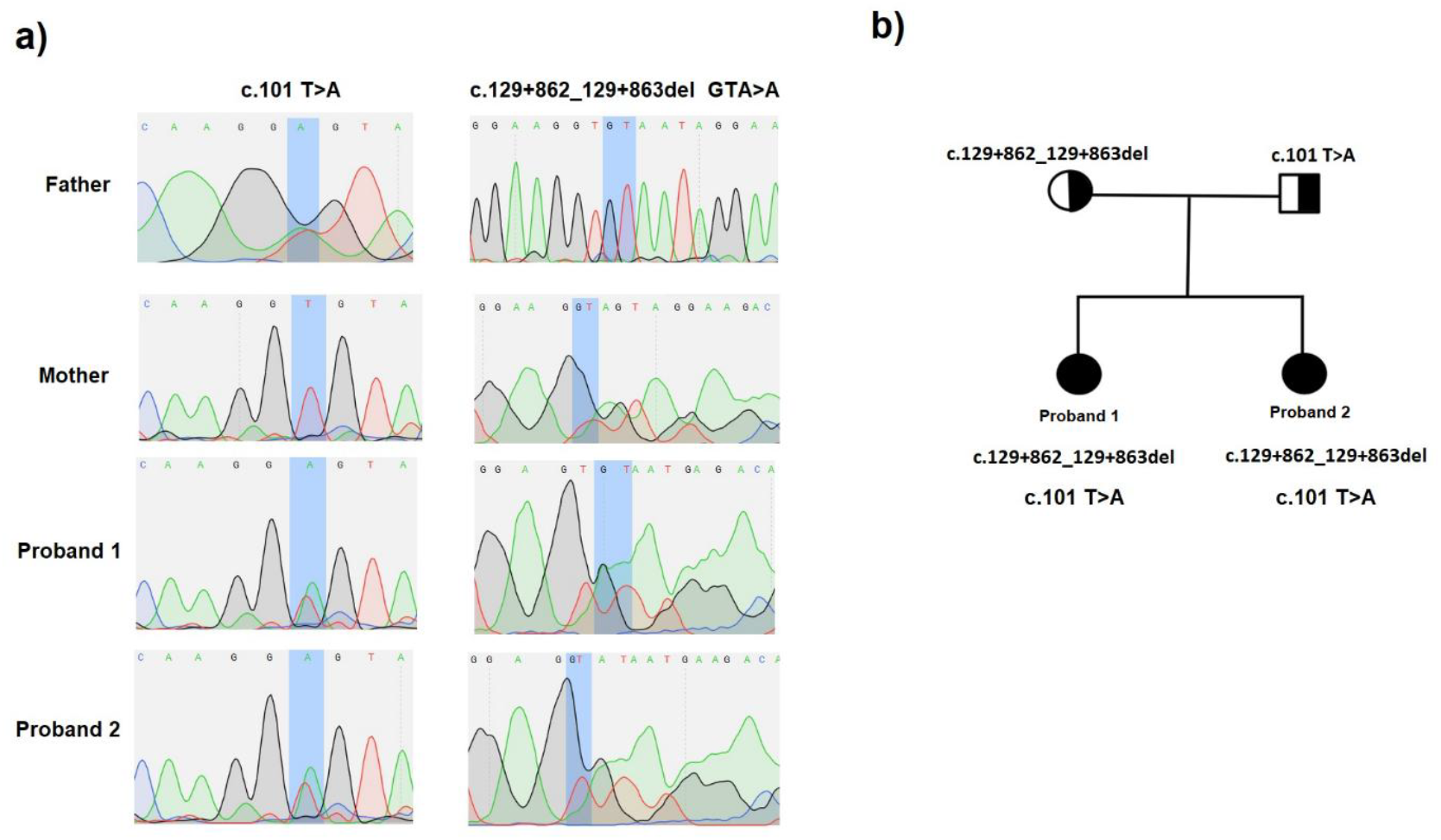
*CYB5A* variants and segregation in the family. (a) Sanger sequencing chromatograms showing the heterozygous *CYB5A* missense variant c.101T>A in the father and both probands, and the heterozygous intronic variant c.129+862_129+863del in the mother and both probands. The probands are compound heterozygotes for c.101T>A and c.129+862_129+863del. Variant positions are indicated by blue shading. (b) Pedigree of the family demonstrating autosomal recessive inheritance of *CYB5A* variants, with the father carrying c.101T>A, the mother carrying c.129+862_129+863del, and both affected siblings carrying both variants.

### RNA splicing assay results

RT-PCR analysis demonstrated distinct amplicon sizes for the pSPL3-*ABCC8*-9-10 splicing control, the *CYB5A* wild-type construct, and the *CYB5A* construct carrying the c.129+862_129+863del intronic deletion (Figure 2a). Sanger sequencing of all three RT-PCR products confirmed the expected splicing patterns. In the pSPL3-*ABCC8*-9-10 control, exon 9 spliced correctly to exon 10 with complete removal of the intervening intron (Figure 2b). In the *CYB5A* wild-type construct, *CYB5A* exon 1 spliced directly to exon 10 within the vector backbone, again demonstrating normal intron removal (Figure 2c). In contrast, the *CYB5A* construct containing the c.129+862_129+863del variant showed aberrant splicing, with exon 1 joined to a retained intronic fragment that formed a pseudo-exon, followed by splicing to exon 10 (Figure 2d). Inclusion of this pseudo-exon introduced a premature stop codon, predicted to generate a truncated CYB5A protein of approximately 50 amino acids.

**Figure 2:**
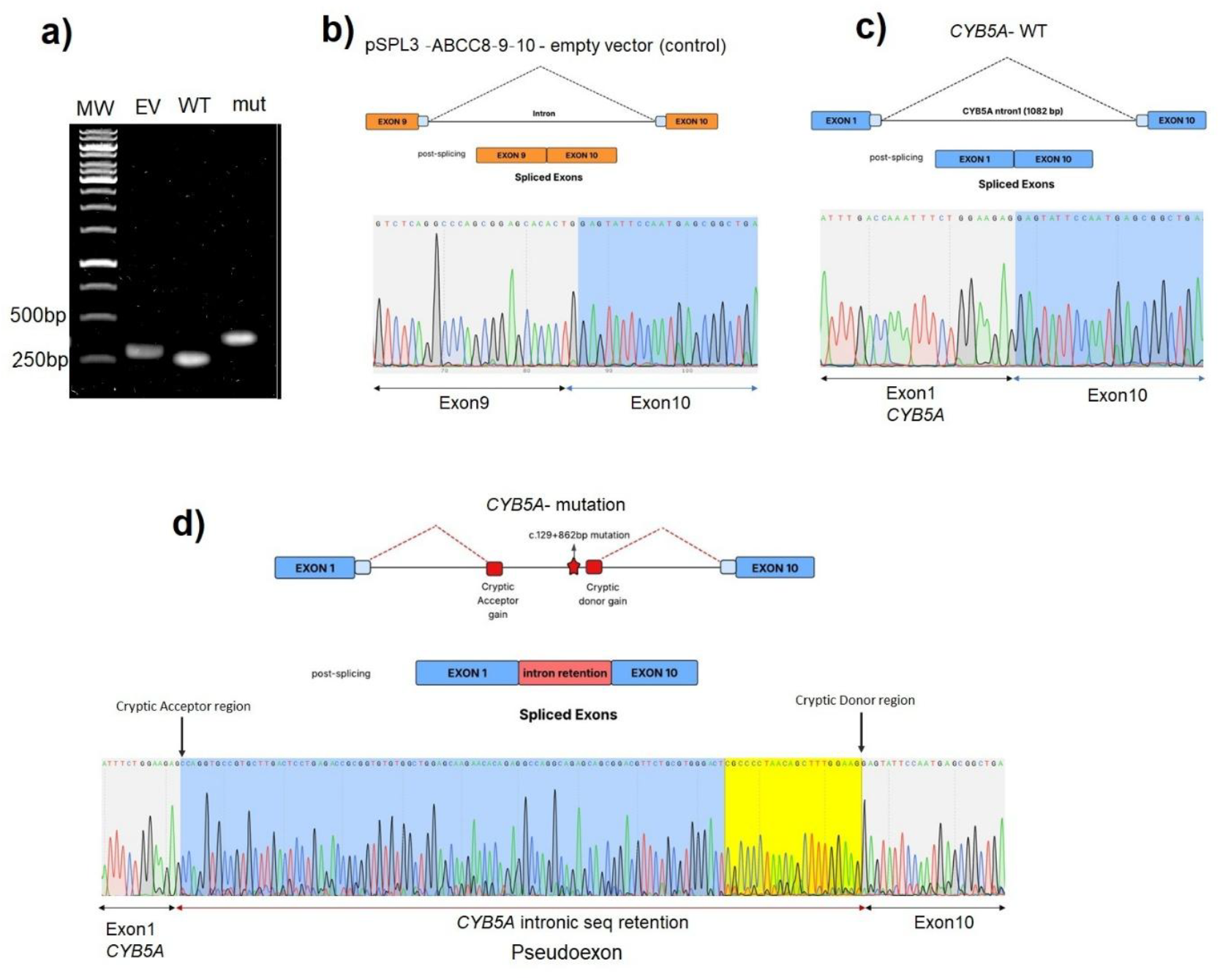
Minigene splicing assay demonstrating aberrant *CYB5A* splicing caused by the intronic variant. (a) RT-PCR analysis of pSPL3-*ABCC8*-9-10 minigene constructs showing distinct band patterns for empty vector (EV), wild-type (WT), and mutant (Mut) *CYB5A* inserts. The mutant construct produces an additional larger amplicon consistent with intron retention or pseudo-exon inclusion. (b) Control pSPL3-*ABCC8*-9-10 splicing assay confirming correct exon– exon junction formation in the empty vector backbone. (c) Sequencing of the WT *CYB5A* minigene transcript demonstrating normal splicing between exon 1 and exon 10. (d) Sequencing of the mutant *CYB5A* minigene transcript showing activation of cryptic acceptor and donor sites, leading to retention of intronic sequence and pseudo-exon formation. The retained intronic region and cryptic splice sites are highlighted.

### CYB5A protein Structure

CYB5A is located on chromosome 18 (chr18:71,920,531–71,958,276) and encodes three protein-coding transcripts, of which the longest isoform (ENST00000340533.4) produces a 134aa protein. The CYB5A protein contains a well-defined cytochrome b5 heme-binding domain spanning amino acids 9–82, encoded by exons 1 and 2. This domain has been experimentally resolved by NMR and represents the critical functional core required for electron transfer activity (25). AlphaMissense (26) analysis highlights this region as highly constrained, with substitutions across the heme-binding domain broadly predicted to be damaging or likely pathogenic. The missense variant identified in our patients, p.Val34Glu, lies within this essential domain, and AlphaMissense classifies substitutions at this position as deleterious, supporting its functional relevance and contribution to disease.

To evaluate the structural consequences of the CYB5A p.Val34Glu substitution, we performed in silico protein modelling (Figure 3). AlphaMissense pathogenicity mapping demonstrated that Val34 lies within a highly constrained region of the heme-binding domain, with a strong predicted deleterious effect (Figure 3a). Structural domain visualization confirmed that Val34 is positioned on the surface of the heme-binding fold, adjacent to residues critical for electron transfer to CYP17A1 (Figure 3b). Comparative modelling in ChimeraX (27) showed that replacement of the non-polar valine with a negatively charged glutamic acid introduces a bulkier, acidic side chain that disrupts local packing and alters the electrostatic environment of the heme-binding pocket (Figure 3c). These structural changes are consistent with impaired CYB5A stability and reduced electron-donor function.

**Figure 3.**
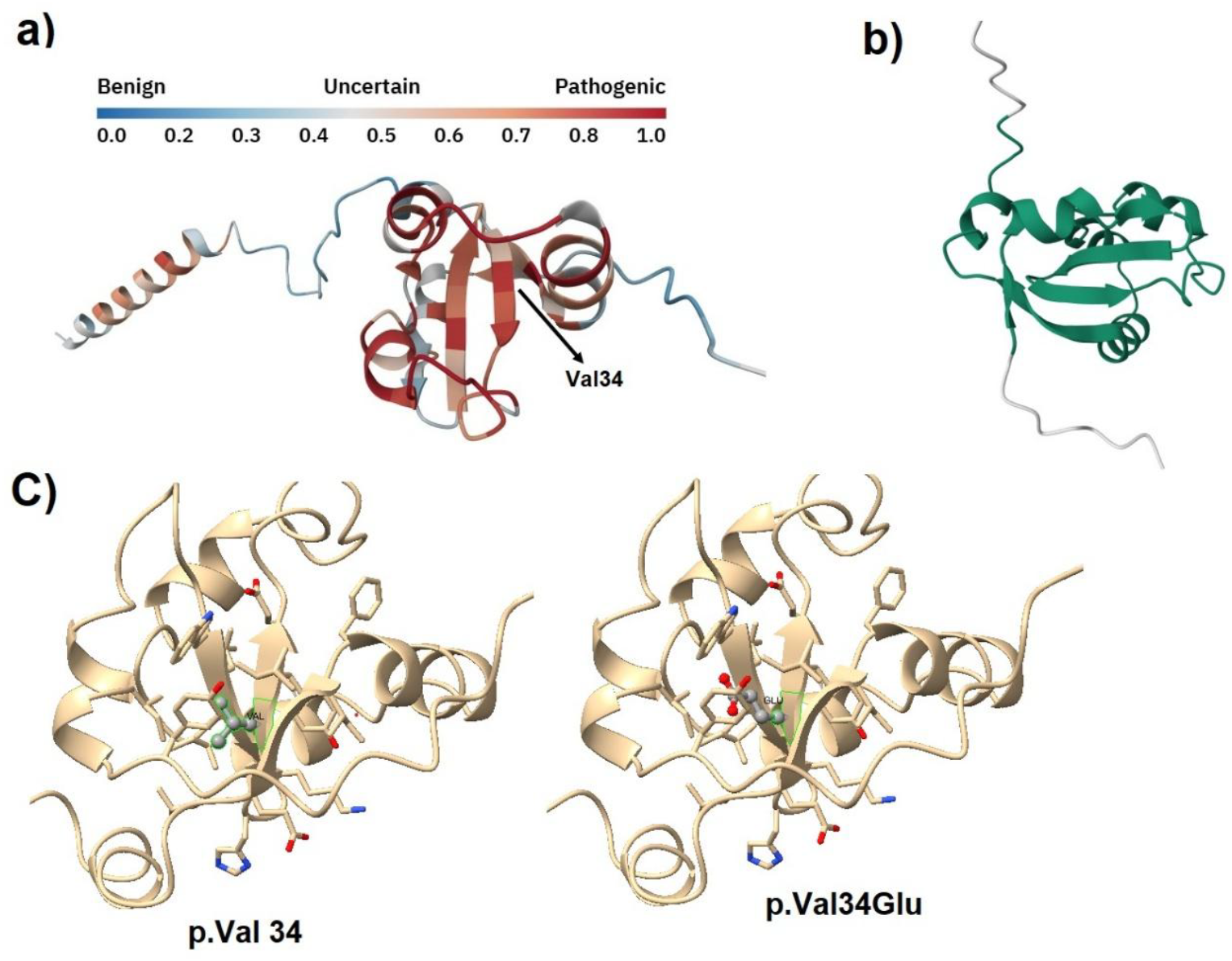
Structural impact of the CYB5A p.Val34Glu variant. a) AlphaMissense pathogenicity heat-map projected onto the CYB5A protein structure, highlighting residue Val34 within the highly constrained heme-binding domain. Warmer colors indicate higher predicted pathogenicity. b) The green region representation of the CYB5A heme-binding domain showing the overall fold and spatial position of the conserved region. c) Structural comparison of the wild-type Val34 residue and the mutant Glu34 modeled in ChimeraX. The substitution introduces a negatively charged side chain at position 34, altering local interactions within the heme-binding domain.

### Variant Classification

Both *CYB5A* variants were classified using ACMG/AMP criteria. The missense variant NM_148923 c.101T>A (p.Val34Glu) locating in a functional cytochrome b5 heme-binding (Cyt-b5) domain, was inherited from the father and lies within a highly conserved region. In silico tools predicted a deleterious effect (REVEL score=0.931, AlphaMissense score = 0.93, CADD score=32), and the variant was absent from gnomAD v4.1.0 and ClinVar. Both affected siblings inherited this missense variant in compound heterozygosity with the c.129+862_129+863del variant, contributing to the segregation evidence with disease. Based on the ACMG/AMP guideline, the variant was classified as Likely pathogenic (PM2_Supporting, PP3, PM1, PP1, PM3_Supporting).

The maternally inherited deep intronic variant c.129+862_129+863del was also absent from population databases (PM2_Supporting) and predicted to create a cryptic donor site and a cryptic acceptor site by SpliceAI. Functional minigene splicing assays demonstrated a 113bp pseudo-exon between exon 1 and exon 2, resulting to an introduction of a premature stop codon after seven aberrant amino acids, predicting a truncated 50aa protein. Nonsense mediated decay is likely to occur, leading to null protein. *CYB5A* variants cause disease via loss of function mechanism. Taken together, the deep intronic variant was classified as Pathogenic (PVS1, PM2_Supp, PM3_Supp).

In conclusion, we identified the two variants as a compound heterozygous likely pathogenic and pathogenic variants in *CYB5A*, confirming a genetic diagnosis for both affected siblings with 46,XY DSD.

## Discussion

CYB5A is an essential cofactor in human steroidogenesis, acting as a selective enhancer of CYP17A1 17,20-lyase activity, the key step required for DHEA and downstream androgen production(11, 14, 28). Disruption of CYB5A function therefore leads to impaired androgen biosynthesis and a hormonal profile consistent with the phenotype observed in our patients. Hormone profiling in the affected siblings revealed a pattern of low testosterone, low estradiol, and normal 17-hydroxyprogesterone levels, along with prepubertal gonadotropin levels. This profile indicates reduced sex-steroid synthesis, which is typical in disorders involving 17,20-lyase activity. While elevated 17 OHP is usually seen in 21-hydroxylase deficiency, cases related to CYB5A show that 17 OHP levels in isolated 17,20-lyase deficiency can vary and often stay within normal ranges, especially in prepubertal children. This occurs because 17-OHP is not the main substrate for the 17,20-lyase reaction and can be redirected into other metabolic pathways, preventing its buildup (28).

The central role of CYB5A in this pathway explains why even subtle alterations in its structure or expression can produce significant disorders of sex development. To date, only three CYB5A variants have been reported in ClinVar as pathogenic or likely pathogenic, reflecting the rarity of CYB5A-related disease. These include a homozygous splice-site variant c.130-2A>G (14, 29), a homozygous missense variant c.131A>T (p.H44L), supported by functional assays and classified as likely pathogenic (30); and a homozygous nonsense variant c.81G>A (p.W27*) (31). Our variants represent the fourth report of pathogenic and likely pathogenic *CYB5A* variants validated by functional assay, expanding the mutational spectrum of this gene. Importantly, this is the first report of compound heterozygous variants in *CYB5A*, demonstrating that pathogenicity can arise from two mechanistically distinct alleles, one affecting protein structure and the other disrupting RNA splicing. This finding also underscores the diagnostic value of WGS. Given the patients’ phenotype, clinicians would typically order WES or a targeted panel sequencing. However, identifying only a single heterozygous missense variant in *CYB5A* would not explain the condition, as the gene follows autosomal recessive inheritance. Our study shows that a deep intronic variant located 861 bp downstream of exon 1 likely disrupted splicing during gonadal development and contribute to disease, and such variants are invisible to WES and most clinical panels.

Therefore, the functional consequences of both variants converge on loss of CYB5A activity. The p.Val34Glu missense variant lies within the heme-binding domain (aa 9–82), a region essential for electron transfer to CYP17A1 (32, 33). Structural prediction tools, including AlphaMissense, classify substitutions at this position as deleterious, consistent with destabilization of the domain. In contrast, the deep intronic deletion activates a cryptic splice site, resulting in pseudo-exon inclusion and introduction of a premature stop codon. The predicted truncated protein lacks the entire heme-binding domain, rendering it non-functional. These complementary mechanisms structural disruption and aberrant splicing, provide a coherent explanation for the biochemical and clinical phenotype.

Our findings also highlight the broader importance of evaluating non-coding regions in patients with unexplained steroidogenic defects. As WGS becomes more widely adopted, deep intronic variants are increasingly recognized as contributors to Mendelian disease. Functional assays such as minigene systems remain essential for validating their impact, particularly when computational predictions alone are insufficient for ACMG classification. Clinically, establishing the molecular diagnosis confirms autosomal recessive inheritance, enables accurate carrier testing, and provides clarity for the family. More broadly, this study reinforces that defects in electron-transfer cofactors, not only in steroidogenic enzymes can lead to disorders of sex development. CYB5A should therefore be considered in cases with isolated 17,20-lyase deficiency, even when coding regions appear normal.

In summary, this work identifies the first compound heterozygous pathogenic variants in *CYB5A*, expands the known mutational landscape of the gene, and demonstrates that a deep intronic deletion can profoundly alter splicing. By integrating WGS, structural prediction, and functional validation, we provide new insights into the essential role of *CYB5A* in human steroidogenesis and highlight the importance of comprehensive variant evaluation in the diagnosis of DSD.

## Acknowledgements

The authors would like to thank the patients and their family for their participation in this study and for providing blood samples.

## Research Funding

The work was funded by NHMRC Idea grant 2002426

## Author contributions

All authors have accepted responsibility for the entire content of this manuscript and approved its submission.

## Data Availability

All data produced in this study are available from the corresponding author upon reasonable request.

